# Silver diamine fluoride, atraumatic restorations, and oral health-related quality of life - Results from the *CariedAway* cluster randomized trial

**DOI:** 10.1101/2021.04.19.21255478

**Authors:** Ryan Richard Ruff, Tamarinda J. Barry Godín, Topaz Murray Small, Richard Niederman

**Affiliations:** New York University College of Dentistry, Department of Epidemiology

## Abstract

**Objective:** Silver diamine fluoride (SDF) is a non-surgical treatment for the arrest and prevention of dental caries that results in irreversible black staining of dental decay. The objective of this study was to evaluate the impact of SDF treatment on oral health-related quality of life (OHRQoL) relative to a standard package of glass ionomer sealants and atraumatic restorative treatment.

**Methods:** *CariedAway* is a pragmatic, longitudinal, cluster-randomized non-inferiority trial of non-surgical interventions for caries. Secondary study outcomes included OHRQoL and academic performance. Oral health-related quality of life was measured at each study visit using the Child Oral Health Impact Profile. Change in OHRQoL was assessed using linear regression and non-inferiority was determined using t-tests.

**Results:** Untreated decay at baseline was associated with signifcantly worse ORHQoL and treatment in both groups resulted in incremental improvement. Quality of life in children receiving silver diamine fluoride was non-inferior to those receiving sealants and ART at least six months post-treatment. Additionally, change in OHRQoL did not depend on the severity of baseline decay.

**Conclusions:** ORHQoL is related to untreated dental caries, however no appreciable change was observed following SDF treatment relative to standard preventive therapies.

## 1. Introduction

Dental caries is the most prevalent childhood disease in the world (Bernabe et al., 2020), found across all age groups and most prominent among low-income populations (Frencken et al., 2017). Untreated caries has been shown to develop into pain and systemic infection, potentially resulting in functional and/or psychosocial impairment (Mathur and Dhillon, 2018). Much of the disproportionate burden of disease amongst vulnerable groups, such as low-income and minority populations, is due to lower accessibility and utilization of traditional dental services (Dye et al., 2012; Griffin et al., 2016; Treadwell, 2017). As a result, the use of non-surgical treatments such as silver diamine fluoride (SDF) is increasing. Silver diamine fluoride is a noninvasive method to prevent and arrest caries that can be efficiently applied in community settings (Contreras et al., 2017; Oliveira et al., 2019; Wierichs and Meyer-Lueckel, 2015), but results in permanent black staining of dental decay.

Oral health-related quality of life (OHRQoL) is a multidimensional construct consisting of subjective evaluations of oral health, functional well-being, emotional well-being, satisfaction with care, and sense of self (Ruff et al., 2017). Caries may have a negative impact in oral health-related quality of life in preschool children (Nora et al., 2018), children aged 3-12 years and adults (Aimee et al., 2017; Haag et al., 2017; Moghaddam et al., 2020). High caries experience (Chaffee et al., 2017) and untreated caries (Fernandes et al., 2017) both exhibit reduced OHRQoL, regardless of measurement used (Arrow, 2017; García-Pérez et al., 2017). Despite a high oral disease burden (Ahluwalia and Sadowsky, 2003), research on quality of life and caries in black and Hispanic/Latino populations is limited (Broder et al., 2000; Southward et al., 2008); evidence on silver diamine fluoride and quality of life presents conflicting results with treatment shown to either improve or have no effect on OHRQoL in children (Duangthip et al., 2019; Jiang et al., 2020, 2019; Rodrigues et al., 2020; Sihra et al., 2020; Vollu et al., 2019); and the impact on OHRQoL comparing SDF to atraumatic restorative treatment is unclear (Rodrigues et al., 2020; Vollu et al., 2019).

*CariedAway* is an active randomized controlled trial of non-surgical interventions for the prevention and treatment of caries in children aged 5-13 years (Ruff and Niederman, 2018), specifically silver diamine fluoride, sealants, and atraumatic restorative treatment. The *CariedAway* study also aims to evaluate the effects of treatment on quality of life, academic performance, and school attendance. The objectives of this paper are to assess (1) the associations between oral health-related quality of life and dental caries and (2) the short term effects of non-surgical treatment for caries on oral health-related quality of life.

## 2. Methods

Ethical approval for the *CariedAway* clinical trial was obtained from the New York University School of Medicine Institutional Review Board (i17-00578). A previously published trial protocol contains additional study-related information (Ruff and Niederman, 2018) and the trial is registered at www.clinicaltrials.gov (#NCT03442309). Preliminary clinical results are forthcoming.

### 2.1. Design

*CariedAway* is a longitudinal, cluster-randomized, single-blind, pragmatic trial with the primary objective of evaluating the non-inferiority of non-surgical treatments for dental caries. Any school in New York City with a student population of at least 80% receiving free or reduced lunch and at least 50% Hispanic/Latino or black was eligible to participate in the study. All children in enrolled schools were provided informed consent, there were no inclusion criteria for child-level enrollment. Any subject with parental informed consent and child assent was randomly assigned to treatment and received care. Treatment was provided in scheduled six-month intervals.

### 2.2. Interventions

Interventions included two separate packages of non-therapeutic treatments for dental caries: a simple combination of fluoride varnish applied to all teeth and silver diamine fluoride applied to all pits and fissures and asymptomatic cavitated lesions of bicuspids and molars, and a complex combination consisting of the same application of fluoride varnish, glass ionomer sealants applied to pits and fissures of bicuspids and molars, and use of atraumatic restorative treatment on all frank asymptomatic cavitated lesions. Caries diagnosis followed the International Caries Detection and Assessment System (ICDAS) adapted criteria and the diagnostic and treatment protocol is previously described (Ruff and Niederman, 2018).

### 2.3. Randomization

Enrolled schools were block randomized in a 1:1 allocation ratio.

### 2.4. Data Collection

At each observation, study clinicians performed a full visual-tactile oral examination and recorded the missing, sound, decayed, or filled status of every tooth surface. Clinicians were standardized and calibrated prior to observing subjects. Following the oral examination and application of treatments, children were asked to participate in answering the COHIP-SF quality of life survey. The clinician would then read each question aloud to subjects who would then note their answers on a provided tablet computer.

### 2.5. Outcomes

Oral health-related quality of life was assessed using the Child Oral Health Impact Profile Short Form (COHIP-SF), consisting of 19 questions assessing oral health, functional well-being, socio-emotional well-being, school-environment, and self-image (Broder et al., 2012; Broder and Wilson-Genderson, 2007). Examiners asked participants each COHIP-SF question (e.g., “Have you in the past three months had pain in your teeth or a toothache”), and children responded by touching an indicator on the tablet or communicating their answer (“Never,” “Almost never,” “Sometimes,” “Fairly often,” “Almost all of the time”) to the examiner. A global quality of life indicator measuring perceived change from the previous observation was also used. For the global QoL question, participants were asked “On a scale of 0 to 100 where 0 is worst and 100 is best, how would you rate your quality of life?” Subjects were then shown a visual scale on a horizontal axis ranging from 0 to 10 in 10-point intervals, with “worst” at the zero point, “medium” at the 50th point, and “best” at the 100th point. Participants then tapped their answer at the point of the scale directly on the tablet. Not all children received the COHIP instrument: fifty percent of participants in the *CariedAway* trial were randomly assigned within each group at baseline to receive the quality of life assessment. Assigned children then received the same QoL instrument at every successive treatment.

### 2.6. Covariates

Demographic information including age, sex, and race/ethnicity were obtained from informed consent documents or school records. A unique identification number maintained by the Office of School Health at the New York City Department Health and Mental Hygiene and New York City Department of Education was similarly used as the patient record number for this study.

### 2.7. Statistical Analysis

Analysis was restricted to only those subjects between the ages of 5 and 13 years at time of observation. Subjects were analyzed using intent to treat: any child who may have switched schools that was randomized to a different treatment arm was analyzed according to his or her original assignment. Baseline descriptive statistics for sociodemographic variables and COHIP scores were computed. The association between dental caries at baseline and initial quality of life was assessed using linear regression. Following baseline analyses, subjects were ordered sequentially by visit date and any child without two completed visits was removed from the analytic sample. Within-group differences for treatment groups used paired samples t-tests. Post-treatment analysis of the caries-QoL association was assessed using linear regression, adjusting for treatment group, baseline COHIP scores, and demographic variables. The intraclass correlation coefficient for subjects nested within schools was estimated using an intercept-only mixed effects multilevel model. All analyses adjusted for the clustering effect of schools. The non-inferiority of SDF therapy compared to sealants/ART on quality of life was determined by calculating the confidence interval for the difference in mean COHIP scores across treatment group. As per the original study protocol, a non-inferiority margin of ten was used.

## 3. Results

1323 subjects completed the quality of life assessment at baseline, 160 of which (12%) completed a second-follow up at least six months following treatment with either silver diamine fluoride or sealants/ART, prior to study suspension due to the impact of the SARS-CoV-2 pandemic. The sample was approximately 53% female and consisted of 39% Hispanic/Latino, 15% black, 4% white, and 1.4% Asian (Table 1). The overall prevalence of any untreated decay on any tooth (deciduous or permanent) was 25.8% with an average per-person number of decayed teeth of 0.45 (SD = 1.1). The average time between treatments was 189 days. The intraclass correlation was 3.26%.

**Table 1:** Baseline sample demographics and clinical outcomes (N=1323)

| Variable | N/Mean | %/SD |
| --- | --- | --- |
| **Sex** |  |  |
| Female | 700 | 52.91 |
| **Ethnicity** |  |  |
| Hispanic | 513 | 38.78 |
| Non-Hispanic | 105 | 7.94 |
| N/A | 705 | 53.29 |
| **Race** |  |  |
| Black | 203 | 15.34 |
| Asian | 19 | 1.44 |
| Multirace | 34 | 2.57 |
| White | 56 | 4.23 |
| N/A | 984 | 74.38 |
| **Clinical Indicators** |  |  |
| Untreated decay | 341 | 25.77 |
| # Decayed teeth | 0.49 | 1.09 (SD) |

Children with untreated decay at baseline, irrespective of treatment assignment, scored significantly worse on oral health-related quality of life (B = 3.76, 95% CI = 2.37, 5.14). Adjusting for differences in race and ethnicity, each decayed tooth was associated with 1.37 point increase in COHIP scores, where higher scores indicate worse OHRQoL (Table 2). There were no baseline differences in OHRQoL by treatment group (B = −0.61, 95% CI = −4.17, 2.94). Across both SDF and sealant/ART groups, average OHRQoL improved following treatment: COHIP scores improved from an average of 16.28 (SD=11.32) in children receiving silver diamine fluoride at baseline to 14.51 (SD=11.83) at follow-up, while those receiving sealants and atraumatic restorative treatments improved from 16.90 (SD=10.26) to 16.38 (SD=11.33). Following at least six months post-treatment, there were no differences in OHRQoL by treatment group (Table 3), adjusting for baseline COHIP scores, the number of decayed teeth, and sociodemographic factors (B = −.083, 95% CI = −3.89, 2.23).

**Table 2:**
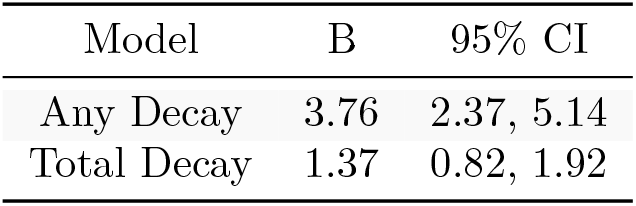
Baseline decay and OHRQoL

**Table 3:** SDF vs ITR for OHRQoL (analytic sample)

| Variable | B | 95% CI |
| --- | --- | --- |
| SDF | -0.83 | -3.90, 2.23 |
| Baseline OHRQoL | 0.68 | 0.55, 0.81 |
| Total decay | 0.34 | -1.91, 2.60 |

<sup>a</sup> Models also adjusted for race and age

Comparisons of the analytic sample at the follow-up visit indicate that self-reported oral health-related quality of life in children receiving silver diamine fluoride is non-inferior to those receiving traditional glass ionomer sealants and ART (mean difference = 1.87, 95% CI = −1.92, 5.66). The point estimate favors silver diamine fluoride however the confidence interval is below the non-inferiority margin. Additionally, there was no significant interaction: no treatment differences were found amongst only subjects with baseline caries.

## 4. Discussion

As arresting and preventive agents for dental caries, atraumatic restorative treatments and glass ionomer sealants are visually imperceptible when compared to traditional composite restorations. In contrast, application of silver diamine fluoride results in permanent black staining of dental decay and superficial staining of the oral mucosa. Notably, perceptions of self are affected by facial aesthetics, being previously observed in adolescents seeking orthodontic treatment and in children with preexisting orofacial anomalies (Phillips and Beal, 2009; Ruff et al., 2016a). As approximately 25% of *CariedAway* participants had untreated decay at baseline with significantly lower OHRQoL than caries-free children, concerns regarding the aesthetic impact of SDF, despite demonstrated clinical and economic benefits (Contreras et al., 2017; Yeung and Argáez, 2017), may be justified.

Oral health-related quality of life slightly improved following treatment with either silver diamine fluoride or sealants/ART, and children receiving SDF exceeded the minimally important difference threshold of the COHIP-SF necessary for patient-centered clinically meaningful change (Ruff et al., 2016b). Results further suggest that OHRQoL in children receiving SDF was non-inferior to those of children receiving ART/sealants. These results are consistent with other studies of SDF in children reporting similar effects when compared to alternative treatments, such as ART, fluoride varnish, or placebo (Duangthip et al., 2019; Jiang et al., 2020, 2019; Sihra et al., 2020; Vollu et al., 2019).

Clinical application of silver diamine fluoride in the *CariedAway* trial does not include anterior teeth; SDF is often applied to posterior teeth in order to avoid impacts on facial aesthetics. In children aged 5-9 years, the global prevalence rate of caries in deciduous teeth exceeds 40% (Bernabe et al., 2020). However, decay most often occurs in the occlusal surface of molars and pre-molars (Demirci et al., 2010), thus a restriction to posterior application will still treat a majority of underlying disease. In previous studies of SDF, caregivers of children with untreated caries were more accepting of the staining effect when applied to posterior lesions, if the child had a history of behavioral issues when treated by a dentist, or if more invasive measures, such as anesthesia, would be required (Seifo et al., 2020). Anterior application of silver diamine fluoride may be acceptable for deciduous teeth due to expected exfoliation, but more aesthetically pleasing alternatives may be required for permanent anterior teeth in adolescents.

The focus of this analysis was on the potential short-term impact of SDF application on oral health-related quality of life, relative to more traditional nontherapeutic caries treatments. It may be the case that the staining effect of SDF, even when confined to posterior teeth, becomes more appreciated with longer rates of follow-up or when children progress into adolescence where facial aesthetics may be of greater concern. Subsequent objectives of the *CariedAway* trial are to assess longitudinal change in OHRQoL and identify latent trajectories of quality of life. Additionally, as overall oral health-related quality of life has been shown to be responsive to the severity of dental caries (Corrêa-Faria et al., 2018), the long-term impact on OHRQoL following treatment with SDF may behave in a similar manner.

Silver diamine fluoride can be applied in significantly less time than atraumatic restorative treatments (Vollu et al., 2019) and does not require the same degree of clinical training, suggesting that SDF is more efficient as a pragmatic treatment for caries. For example, some states authorize registered nurses to provide SDF under the supervision of a licensed general dentist. Additionally, the non-invasive nature of SDF as an arresting agent, combined with its secondary preventive effects, make it an attractive alternative to more traditional non-surgical interventions (Horst and Heima, 2019). Our results suggest that children do not perceive any negative impacts on oral health-related quality of life approximately six months following application. These findings, combined with documented evidence of safety and clinical efficacy, further support the continued use of silver diamine fluoride.

## Data Availability

Data will be available from investigators upon reasonable request at the completion of the trial.

http://www.github.com/ryanruff/

## 5. Supporting Information

### 5.1. Funding

This work was supported in part by an award from the Patient Centered Outcomes Research Institute (#PCS160936724). The content is solely the responsibility of the authors and does not necessarily reflect the official views of the funding organization, New York University, or the NYU College of Dentistry.

## 5.3. Acknowledgements

The authors would like to acknowledge the following members of the *CariedAway* project team: Rachel Whittemore, Nydia Santiago-Galvin, Haley Gibbs, Catherine McGowan, Priyanka Sharma, and those providing clinical care.

